# Universal digital high resolution melt analysis for the diagnosis of bacteremia

**DOI:** 10.1101/2023.09.07.23295215

**Authors:** April Aralar, Tyler Goshia, Nanda Ramchandar, Shelley M. Lawrence, Aparajita Karmakar, Ankit Sharma, Mridu Sinha, David T. Pride, Peiting Kuo, Khrissa Lecrone, Megan Chiu, Karen Mestan, Eniko Sajti, Michelle Vanderpool, Sarah Lazar, Melanie Crabtree, Yordanos Tesfai, Stephanie I. Fraley

**Affiliations:** Department of Bioengineering, University of California, San Diego, La Jolla, CA, USA; Department of Pediatrics, Naval Medical Center San Diego, San Diego, CA, USA; Department of Pediatrics, Division of Infectious Diseases, University of California, San Diego, La Jolla, CA, USA; Department of Pediatrics, Division of Neonatology, The University of Utah, Salt Lake City, UT, USA; MelioLabs, Inc, Santa Clara, CA, USA; Department of Pathology, University of California, San Diego, La Jolla, CA, USA; Department of Pediatrics, Division of Neonatology, University of California, San Diego, La Jolla, CA, USA; Department of Pathology and Laboratory Medicine, Rady Children’s Hospital – San Diego, San Diego, San Diego, CA, USA

## Abstract

Fast and accurate diagnosis of bloodstream infection is necessary to inform treatment decisions for septic patients, who face hourly increases in mortality risk. Blood culture remains the gold standard test but typically requires ∼15 hours to detect the presence of a pathogen. Here, we assess the potential for universal digital high-resolution melt (U-dHRM) analysis to accomplish faster broad-based bacterial detection, load quantification, and species-level identification directly from whole blood. Analytical validation studies demonstrated strong agreement between U-dHRM load measurement and quantitative blood culture, indicating that U-dHRM detection is highly specific to intact organisms. In a pilot clinical study of 21 whole blood samples from pediatric patients undergoing simultaneous blood culture testing, U-dHRM achieved 100% concordance when compared with blood culture and 90.5% concordance when compared with clinical adjudication. Moreover, U-dHRM identified the causative pathogen to the species level in all cases where the organism was represented in the melt curve database. These results were achieved with a 1 mL sample input and sample-to-answer time of 6 hrs. Overall, this pilot study suggests that U-dHRM may be a promising method to address the challenges of quickly and accurately diagnosing a bloodstream infection.

**Universal digital high resolution melt analysis for the diagnosis of bacteremia:** April Aralar, Tyler Goshia, Nanda Ramchandar, Shelley M. Lawrence, Aparajita Karmakar, Ankit Sharma, Mridu Sinha, David Pride, Peiting Kuo, Khrissa Lecrone, Megan Chiu, Karen Mestan, Eniko Sajti, Michelle Vanderpool, Sarah Lazar, Melanie Crabtree, Yordanos Tesfai, Stephanie I. Fraley

## INTRODUCTION

Sepsis is among the most common causes of death in hospitalized patients. One out of every five deaths worldwide are estimated to be due to sepsis-related complications, with 41% occurring in children.^1^ Early detection of the infectious cause is critical for sepsis survival, as every hour the infection goes undiagnosed or inaccurately treated mortality risk rises by 4%.^2–4^ However, rapid diagnosis of bloodstream infection (BSI) has proven to be extremely challenging.^5^ Blood culture remains the gold-standard test despite significant false positive and negative error rates and slow time-to-result, ranging from ∼15 hours to 5 days.^6–12^ Consequently, treatments remain un-targeted, contributing to antimicrobial resistance and opportunistic infections.

Nucleic acid amplification tests (NAATs) have been heralded as the solution to this challenge. However, NAATs are associated with higher rates of bacterial DNA detection compared to bacterial growth in blood culture. They also detect bacterial DNA in the blood of healthy patients, which has constrained their impact and widespread adoption by the medical community. For example, multiplexed PCR detection of microbial DNA in blood suffers from false positives that overestimate the concentration of pathogens in clinical samples.^13–15^ False negatives also arise from low concentrations of pathogens and subsampling errors.^16–18^ Likewise, the off-target interaction of primers with human DNA, which vastly outnumbers bacterial DNA in blood, can contribute to both false-positive and false-negative test results.^19–26^ These limitations extend to commercialized PCR-based strategies such as Iridica, Septifast, and SeptiTest.^7,12^

Sequencing technologies have recently emerged in an attempt to address some of the limitations of more targeted NAATs, but have had limited utility in actual clinical settings.^27–32^ Their primary advantage is broad-based detection, but they detect a wide variety of background DNA of no clinical significance, making the interpretation of their results difficult. ^19,29,30,33^ Recent studies show that they can be helpful in cases of rare pathogens, but add little to no clinical value otherwise.^27,28,34,35^ Additionally, sequencing results are not quantitative, a feature which may help in distinguishing symptomatic infections from colonization or contamination.^28,32^ They are also limited to the send-out format of testing due to their high complexity, batch processing, large format equipment, and expertise required to run and analyze the data.^36–40^ Therefore, they cannot be implemented in typical hospital labs. Most importantly, multiple studies have shown that sequencing take even longer than the average blood culture to report an answer.^27,28^

Towards advancing the field of BSI diagnostics, we have developed an approach called universal digital high-resolution melt analysis (U-dHRM)^41–43^, which conducts universal bacterial amplification in digital PCR (dPCR) followed by digital high resolution melt (dHRM) analysis of DNA amplicons to identify and quantify organisms by their sequence-specific melt curve fingerprints with machine learning (ML)^44,45^. This approach distinguishes itself from standard NAAT HRM-based diagnostics by its potential for unbiased broad-based pathogen identification, absolute quantification of organism(s) load (even in polymicrobial samples), and capability to expand organism identification without assay redesign. To achieve breadth and specificity, U-dHRM relies on probe-free melting of long amplicons covering extensive sequence hypervariability in barcoding genomic regions and uses all the temperature points of multimodal melt curve signatures in a Tm-independent manner to identify organisms.^44,45^ Absolute quantification and single genome analysis is achieved by digital partitioning of genomes into separate reactions for individual melt analysis and quantification.^46,47^ Previous work established proof-of-principal for these functions.^44,45^

Here, we advanced U-dHRM for analytical and clinical performance testing directly from whole blood matrix. Sample preparation techniques that enrich for intact pathogens were integrated upstream of U-dHRM and universal bacterial primers were optimized to limit off-target interactions with human DNA that carries over from blood during extraction. To power reliable ML classification on clinical samples, thousands of digital training curves were generated for 11 clinically relevant pathogens spiked into whole blood. The performance of this advanced U-dHRM workflow was tested on 21 patient samples, where it achieved accurate and fast organism identification and quantification directly from whole blood in 6 hr.

## METHODS

### Whole Blood Sample Preparation

The Molysis Complete 5 kit (Molzym GmbH& Co. KG, Bremen, Germany) was selected for whole blood sample preparation prior to U-dHRM analysis. Molysis is designed to deplete host DNA and enrich for microbial DNA from intact cells. The residual concentration of human DNA following Molysis processing of 1mL whole blood was quantified by dPCR with β-actin gene primers (see PCR Master Mixes section for details) to be 4.67*10^2^-5.67*10^3^ copies/μL in the sample extraction elute (Supplementary Fig. 1). This indicated that human DNA can carryover into a significant proportion of the available partitions of the dPCR chip during U-dHRM analysis and prompted the screening of universal bacterial 16S primers for off-target interactions with human DNA, given their known homology^19^.

### Primer Selection and Internal Amplification Control

Universal primer sequences were identified through literature review and tested in BLAST for alignment to the human genome. Apart from V6R which was used as a control sequence, primer sequences were selected for screening if there was <85% query coverage. As an additional strategy for promoting the detection of intact microbes, primer pairs were required to produce a long amplicon (>600 bp) capable of discriminating against degraded DNA.^48–50^. Amplicons were also required to include the important hypervariable regions (V4 and V6) for differentiation of the greatest number of bacterial species.^51^ Supplementary Table 1 lists the primer pairs selected for screening and their characteristics.

To assess off-target interactions, human genomic DNA was extracted from healthy cord blood using the Wizard Genomic DNA Purification Kit (Promega, Madison, WI) and spiked into qPCR reactions at post-Molysis concentrations (approximately 1 ng/uL DNA based on spectrophotometer reading) with 16S primers. Results were confirmed in dPCR. In qPCR, several primer pairs amplified earlier than others and produced melt curves that were consistent within a primer pair (Supplementary Fig. 2A), suggesting off-target yet reliable interactions. V1F/V9R showed no off-target amplification in qPCR, indicated by the absence of both a cycle threshold (Ct) curve and a melt curve (Supplementary Fig. 2A). Similar results were found in dPCR. Supplementary Fig. 2B shows the amplified partitions from dPCR and the resultant dHRM melt curves for each primer pair. Taller and more consistent melt curves indicate stronger interactions between the primers and the human DNA. The V1F/V9R primer pair produced the smallest number of off-target curves, and they were noisy, short, and inconsistent, indicating weak and rare interactions (Supplementary Fig. 2C, green box). The number of off-target melt curves counted for each primer pair is shown in Supplementary Fig. 2C. Additional testing with no template control (PCR water) and bacterial DNA (*E. coli*) further demonstrated that V1F/V9R has low off-target and high on-target amplification (Supplemental Fig. 3).

V1F/V9R was then confirmed in qPCR to amplify bacterial gDNA extracted from different species representing common bloodborne pathogens in the pediatric population (Supplementary Table 2)^52^. Supplementary Fig. 4A shows the alignment of the primer pair to the 16S gene of each organism, and Supplementary Fig. 4B shows singular, consistent melting curves for each of the bacterial amplicons, indicating that amplification was specific. With the addition of post-Molysis levels of human DNA (Supplementary Fig. 4C), similar melt curves were produced. Their amplitude was slightly lower, suggesting that slight dPCR amplification inhibition occurs in the presence of residual human DNA. These qPCR results verified that V1F/V9R is capable of broad-based bacterial DNA amplification and exclusion of human DNA amplification at concentrations expected following host DNA depletion by Molysis.

Finally, the behavior of the V1F/V9R primers were evaluated in dPCR for contrived bacteremic blood samples pre-processed by Molysis. For these experiments, we also included our previously developed internal amplification control (IAC) that improves quantification in dPCR ^53^. The IAC generates an amplicon that melts at a lower temperature than bacterial amplicons in dHRM. We verified that the IAC assay components do not interact with V1F/V9R by generating blood matrix spike (MS) samples with *E. coli* or no template control (NTC) samples with sterile PBS, processing them with Molysis, and analyzing them by U-dHRM. Supplementary Fig. 5A shows that for the *E. coli* spiked sample, both the IAC and *E. coli* melt curves or the IAC melt curves alone or no melt curves are produced per partition at expected levels. Supplementary Fig. 5B shows that the NTC sample only generated melt curves for the IAC or were negative for amplification. IAC negative reactions are excluded during concentration calculations to improve quantification.^53^

### PCR Master Mixes

All PCRs were performed using a 15-μL total reaction volume. All PCR reactions with the exception of those conducted for identity verification by sequencing and human DNA quantification experiments consisted of 1× Phusion GC PCR buffer (Thermo Scientific, Waltham, MA), 2× ROX dye (Bio-Rad, Hercules, CA), 0.02 μM IAC primers, 0.1 μM IAC template, 0.1 μM each bacterial primer (IDT), 2.5× EvaGreen (Biotium, Fremont, CA), 0.2LmM deoxynucleoside triphosphate (dNTP) (Invitrogen, Carlsbad, CA), 0.02 U/μL Phusion polymerase (New England Biolabs, Ipswich, MA), 3 μL of genomic DNA dilution, and ultrapure water (Quality Biological, Gaithersburg, MD). Identification verification and human DNA quantification experiments were performed in qPCR without the IAC template and bacterial primers in the master mix.

The following β-actin primer sequences were used ^54^: Forward 5’-CGGCCTTGGAGTGTGTATTAAGTA-3’, Reverse 5’-TGCAAAGAACACGGCTAAGTGT-3’.

The internal control template and primer were used at the concentrations determined previously to promote amplification in a maximum number of wells without outcompeting the amplification of low-level bacterial targets.^53^ The IAC template sequence is as follows ^53^: 5’-CCATAGACGTAGCAACGATCGTGAGGTAGTAGATTGTATAGTTGATGCAAGGACTA TCCACTCAC-3’. The IAC was linearly amplified using only a forward primer 5’-CGATCGTTGCTACGTCTATGG-3’.

### PCR and HRM Cycling Conditions

The qPCR experiments were performed on a Bio-Rad CFX 96 (Bio-Rad, Hercules, CA). U-dHRM thermocycling was performed on a ProFlex 2 x Flat Block Thermal Cycler (Applied Biosystems, Waltham, MA). For β-actin amplification, thermocycling for both dPCR and qPCR proceeded as follows: hold at 98°C for 30 s, followed by 70 cycles of 98°C for 10 s, 65.2°C for 30 s, and 72°C for 45 s. For all other reactions, thermocycling for both dPCR and qPCR proceeded as follows: hold at 98°C for 30 s, followed by 70 cycles of 98°C for 10 s, 62°C for 30 s, and 72°C for 45 s. qPCR amplification was followed by a melt cycle of 95°C for 15 s, 45°C for 60 s, and 96°C for 5 s). dPCR amplification was followed by a melt cycle on a Melio MeltRead™ Platform dHRM heating device and simultaneously imaged on a custom Olympus microscope setup as previously described.^46,55^

### Generation of Spiked and Control Blood Samples

Eleven bacterial species were obtained from either the American Type Culture Collection (ATCC), from the Pride Lab at the University of California at San Diego (UCSD), or from the Microbiology Lab at Rady Children’s Hospital, San Diego (RCHSD). The bacteria were cultured in liquid media according to ATCC guidelines. To verify organism identities, genomic DNA from each species was extracted using the Wizard Genomic DNA purification kit (Promega Corporation, Madison, WI). The DNA concentration was measured by biospectrophotometer and diluted in one 10-fold serial dilution before undergoing qPCR amplification with 16S primers. DNA products were then sent for Sanger sequencing.

#### Melt Curve Database

To generate the melt curve database for these organisms, each bacterial species was cultured in liquid culture in the ATCC recommended broth overnight, and 1 mL of freshly prepared bacterial suspension was centrifuged and resuspended in sterile PBS. The turbidity was adjusted to OD_600_ = 1, measured on an biospectrophotometer. Each species of bacteria was spiked into 2 mL of healthy human cord blood at a ratio of 1:10 bacterial suspension to blood. The sample was then split, with 1 mL cultured on agar plates to verify bacterial growth while the other mL went through the Molysis Complete 5 Microbial DNA isolation protocol (Molzym, Bremen, Germany). DNA products were amplified by qPCR and identity verified by Sanger sequencing. Following identity verification, DNA samples were diluted in one ten-fold dilution and analyzed by U-dHRM as described above. This spiking and extraction process was performed twice for each organism to encompass 2 biological replicates, and technical replicates were created from each extraction. Each organism had a total of 8 chips analyzed to create the bacterial melt curve database. The melt curves from each organism were combined and clustered according to the ML algorithm described below and in the supplemental materials. Representative melt curves were generated by the ML algorithm, aligned by their highest peaks, and averaged (see Supplementary Methods).

#### Analytical Validation Studies

Mock samples for analytical validation studies were generated in a similar manner. Briefly, *E. coli* was cultured in LB broth overnight, and 1 mL of freshly prepared bacterial suspension was centrifuged and resuspended in sterile PBS. The turbidity was adjusted to OD_600_ = 0.5, measured on an Eppendorf Biospectrophotometer. Six 10-fold serial dilutions were conducted to achieve concentrations down to 0.1 CFU/mL. Bacteria from each concentration were spiked into 2 mL of healthy human cord blood to achieve the final concentrations of 10k, 1k, 100, 10, 1 and 0.1 bacterial cells/mL of blood. Concurrently, an NTC sample was prepared by adding sterile PBS to the blood at the same volume as the bacterial spike. One mL of each sample was split off and underwent Molysis Complete 5 Microbial DNA isolation and quantification by U-dHRM. The other mL of blood underwent quantitative blood culture (QBC) by plating 100 μL of blood on 10 agar plates to verify bacterial growth and quantity CFU.

Elution blank samples were also prepared to assess the level of background in the reagents and disposables. These were generated following Molzym’s standard negative control test procedure, which consists of processing 1mL of SU buffer (included in the Molysis Complete 5 Kit) through the standard Molysis microbial DNA isolation protocol. Three elution blanks were processed and each was analyzed by U-dHRM across 6 chips following the same procedure as was used for the spiked matrix samples.

### Clinical Sample Testing Procedure

For all scavenged whole blood samples, 1 mL of blood was processed by the Molysis Complete 5 kit (Molzym, Bremen, Germany). Immediately afterwards, the DNA elute was processed concurrently by qPCR detection and U-dHRM quantification using the PCR master mix formulas for bacterial amplification and the PCR protocols described above. Three reactions were run for qPCR testing and three reactions were performed in U-dHRM for each sample. For each method, 9 μL total was sampled out of the 100 μL elute.

### Clinical Samples Scavenging Protocol and Inclusion/Exclusion Criteria

The study protocol was reviewed and approved by the Institutional Review Board (IRB) and Ethics Committee (No. 191392) of the University of California, San Diego (UCSD) and Rady Children’s Hospital in San Diego (RCHSD). A materials transfer agreement (MTA) between RCH and UCSD was executed to enable sample transfer. The age ranges of samples used for this study are shown in Supplementary Table 3. In total, 21 samples were scavenged that met inclusion/exclusion criteria as defined below. Among both positive and negative scavenged CBC samples, 42.9% of the samples were female and 57.1% were male. Racially, 19% of the samples were Asian, 9.5% were black or African American, 28.6% were Hispanic/Latino/Latinx, and 42.9% were White.

#### Positives

Remnant CBC samples were scavenged from the RCHSD hematology lab. CBCs were tagged for scavenging if a blood culture result from the same patient was flagged positive in RCHSD’s clinical microbiology lab. Once a CBC was tagged for collection, an honest broker de-identified the sample and logged sample characteristics. The sample was then transferred to UCSD and processed by U-dHRM the same day. Scavenged CBCs were excluded if they did not meet the following criteria:

(1) The CBC and positive blood culture sample were drawn at the same time (within 1 minute).
(2) No antibiotics were administered within the same hospital visit prior to collection.
(3) At least one milliliter was scavenged.

Seven CBC samples met these criteria during the collection timeframe. It is important to note that these blood samples were first held at room temperature prior to CBC processing and were then transferred to refrigerated storage at 4L in the clinical microbiology lab, where there was a minimum 24-hour hold time until the sample was released to be scavenged. Most pathogenic bacteria do not grow under such conditions^56,57^, so it is unlikely that this hold time would substantially increase the bacterial load that U-dHRM quantifies. However, it is possible that some bacterial cell death occurs during this time.

To assess this possibility, we conducted mock sample aging studies. Briefly, *E. coli* was cultured in LB broth overnight, and 1 mL of freshly prepared bacterial suspension was centrifuged and resuspended in sterile PBS. The turbidity was adjusted to OD_600_ = 0.5, measured on a biospectrophotometer. Ten-fold serial dilutions were conducted and bacteria from two concentrations were spiked into 2 mL of healthy human cord blood to achieve the final concentrations of 10,000 and 1,000 bacterial cells/mL of blood. The 2mL sample was then split: 1mL was immediately processed and 1mL was stored at 4°C for 24 hours before being processed by the Molysis DNA extraction method and subsequent U-dHRM analysis. The measured concentrations of bacteria were compared before and after refrigeration. This experiment was conducted 3 times. A slight but insignificant drop in was detected after refrigeration at both spike concentrations, as shown in Supplemental Fig. 6. While this is not an exhaustive sample aging study, it suggests that the number of organisms detected in the CBC samples may not be strongly impacted by refrigerated storage time. It is important to note that our molecular detection approach only requires organisms to be intact such that their gDNA is protected from degradation during Molysis sample preparation. Viability or culturability is not a requirement for detection by U-dHRM.

#### Negatives

Within the same IRB protocol described above, remnant CBCs matched to blood culture negative samples were scavenged from RCHSD. CBCs were tagged for scavenging if a blood culture result from the same patient resulted as negative in RCHSD’s clinical microbiology lab. Once a CBC was tagged for collection, an honest broker de-identified the sample and logged sample characteristics. The sample was then transferred to UCSD and processed by U-dHRM the same day. During the collection timeframe, 14 negative samples were scavenged. The majority of these were found to have had antibiotic exposure at the time of blood draw. Therefore, we amended our exclusion criteria for negatives to ensure sufficient sample size. Scavenged negative CBCs were excluded if they did not meet the following criteria:

(1) The CBC and blood culture negative sample were drawn at the same time (within 1 minute)
(2) At least one milliliter was scavenged

### U-dHRM Data Curation, Preprocessing, and Melt Curve Classification

A detailed description of the melt curve analysis methods, including the Python packages used, can be found in the Supplementary Methods. Briefly, we employed an optimized image processing, melt curve preprocessing, and machine learning (ML) pipeline for extracting, analyzing, and classifying curves. Fluorescence data extraction from dPCR chips imaged during the dHRM heating process was performed using the protocol detailed in ^53^, and with appropriate temperature-to-time mapping, each resulting raw melt curve was converted to a time series (TS). These fluorescence TS were converted to derivative TS and filtered using several peak-based criteria. Finally, TS smoothing, cropping, and normalizations were performed to preprocess the data into a format appropriate for the downstream ML algorithms.

A two-step classification process was developed for derivative TS classification. Since the melt curves have significant variations and noise within and between chips, K-means clustering was used as a first step to extract the key clusters of variations along with respective cluster centers, which are more robust compressed signals. In the second step, a k-Nearest Neighbour (kNN) classifier was used to classify a test curve by comparing its distance from the cluster centers from the first step. Both the Dynamic Time Warping (DTW) distance, which can take y-axis scaling or x-axis shifting variations into account, and the Euclidean distance are measured, and compared for agreement. If the closest organism call matches by both DTW and Euclidean distance, then the test cluster is confidently called as that respective organism. If the DTW and Euclidean calls do not match, the cluster is considered “low confidence”. Low confidence called clusters can stem from noisy signals, or from novel curves that may not be represented in the database yet.

Classification accuracy was assessed for the database of organism melt curves by dividing the database curves into training and test sets and conducting cross validation experiments. For patient samples, the extracted TS are first clustered using DTW-based K-means, and the obtained cluster centers, or representatives, are classified using the kNN-based classifier that was built using the database curves.

### Statistical Analysis

All statistical analyses were performed in Graphpad Prism 5 (Dotmatics, Boston, MA). A one-way analysis of variance (ANOVA) was conducted on the analyzed testing groups (extraction blanks, culture negative matched CBCs, and culture positive matched CBCs) with a Bonferroni posttest to compare all pairs of columns.

## RESULTS

### Digital Melt Curve Database Generation and Classification in Spiked Whole Blood Matrix

Whole blood matrix spike (MS) samples were generated for 11 organisms (Supplementary Table 2), representing some of the most common causes of BSI among pediatric patients^52^, and analyzed by U-dHRM with primers targeting the V1-V9 regions of the bacterial 16S rRNA gene. Fig. 1A shows representative melt curves for each organism, highlighting the clear visual differences between the V1-V9 melt curve fingerprints. Raw curves can be seen in Supplementary Fig. 7. Coagulase-negative staphylococci (CoNS) species (*S. hominis* and *S. epidermidis*) were combined since their treatment does not differ and they are often considered contaminants. A ML algorithm, which relies on agreement between two classification strategies (see Methods section), demonstrated an average of 97% classification accuracy on this dataset, which included >146,000 training curves (Supplementary Table 4). The number of curves (support) that were used in this analysis for each organism is also detailed in Supplementary Table 4. A confusion matrix resulting from leave one out cross validation (LOOCV) experiments is shown in Fig. 1B. Consistent with the classification accuracy score of 97%, the confusion matrix shows that the vast majority of digital melt curves are classified as the correct organism. Misclassifications occurred rarely, mostly in the *Staphylococcus* genus. These results demonstrate the ability of V1-V9 U-dHRM combined with ML to reliably and automatically differentiate organisms in spiked blood samples.

**Figure 1.**
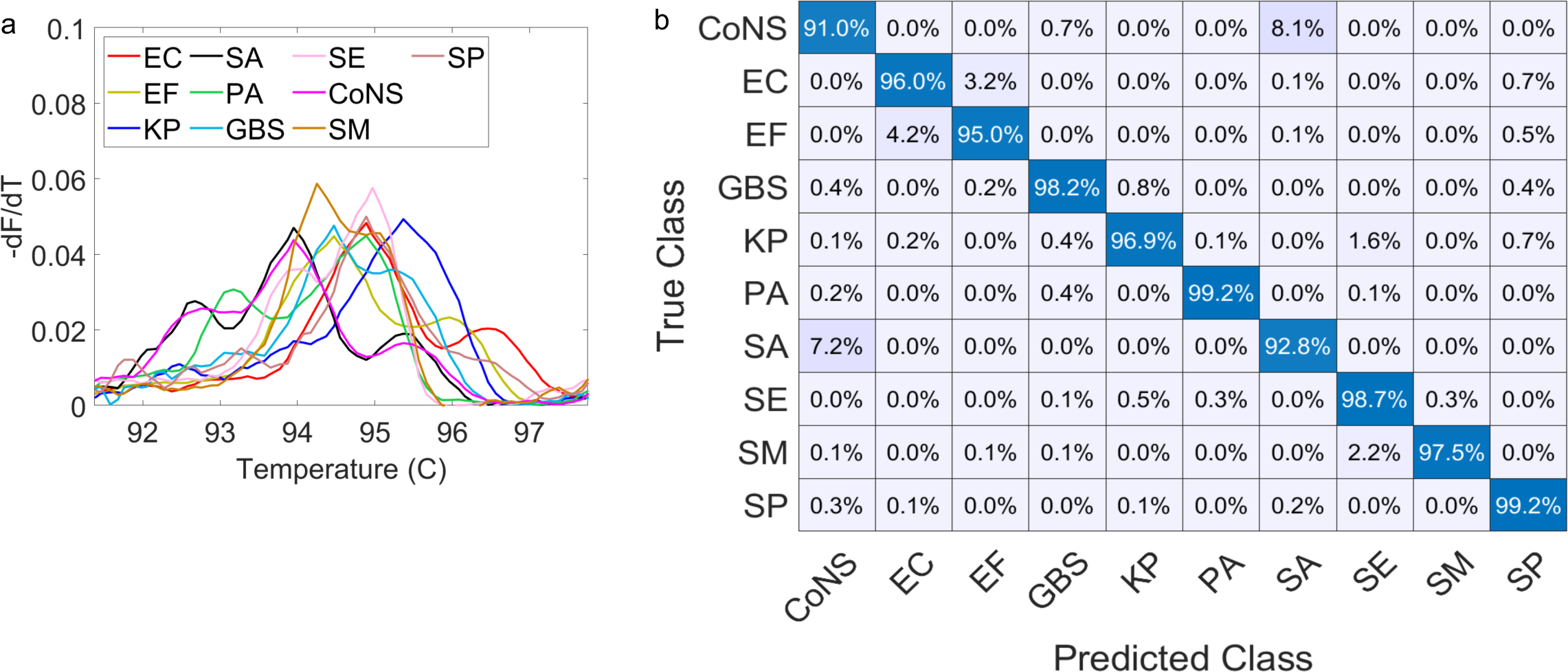
Digital melt curve database creation and algorithm training. Eleven organisms were individually spiked into whole blood, processed with Molysis, and analyzed by U-dHRM with V1F/V9R primers and IAC. A) Plot of representative digital melt curve signature for each organism with the IAC curve region cropped out for visualization purposes. Raw organism curves are shown in Supplementary Fig. 7. B) Confusion matrix showing digital melt curve classification results for the machine learning algorithm.

### Analytical Validation

As a preliminary assessment of quantitative power of U-dHRM, six concentrations of *E. coli* were spiked into a healthy human blood matrix and compared to a no template control (PBS) spiked blood samples. These samples were split in half and underwent paired testing by U-dHRM analysis and quantitative blood culture (QBC) (Fig. 2A). U-dHRM quantification showed excellent agreement with QBC in the concentration range of 10,000-10 CFU/mL, with a Pearson r value of 0.9988 and a p-value of 0.0012 (Fig. 2B). However, at concentrations below 10 CFU/mL, detection variability increased as a result of the extraction elute volume sampled by U-dHRM. From 1mL of blood, extraction elutes 100ul and U-dHRM sampled 9ul of this elute across three chips (3ul each). This results in 1 curve detected per 3 chips equating to 11.1 CFU/mL, and limits quantification below this level. For this reason, when bacteria are spiked at dilutions lower than 10 CFU/mL, U-dHRM quantifies bacteria at 0 CFU/mL (0 curves) or 10 CFU/mL (1 curve). However, for practical reasons, we continued with this format for this pilot study. Extraction blank (EB) samples were tested according to the manufacturer recommended negative control method, which showed a very low level of background originating from the reagents and/or disposables used in the process (Fig. 2B). ML classification of these curves against our database did not identify any matches to the database organisms.

**Figure 2.**
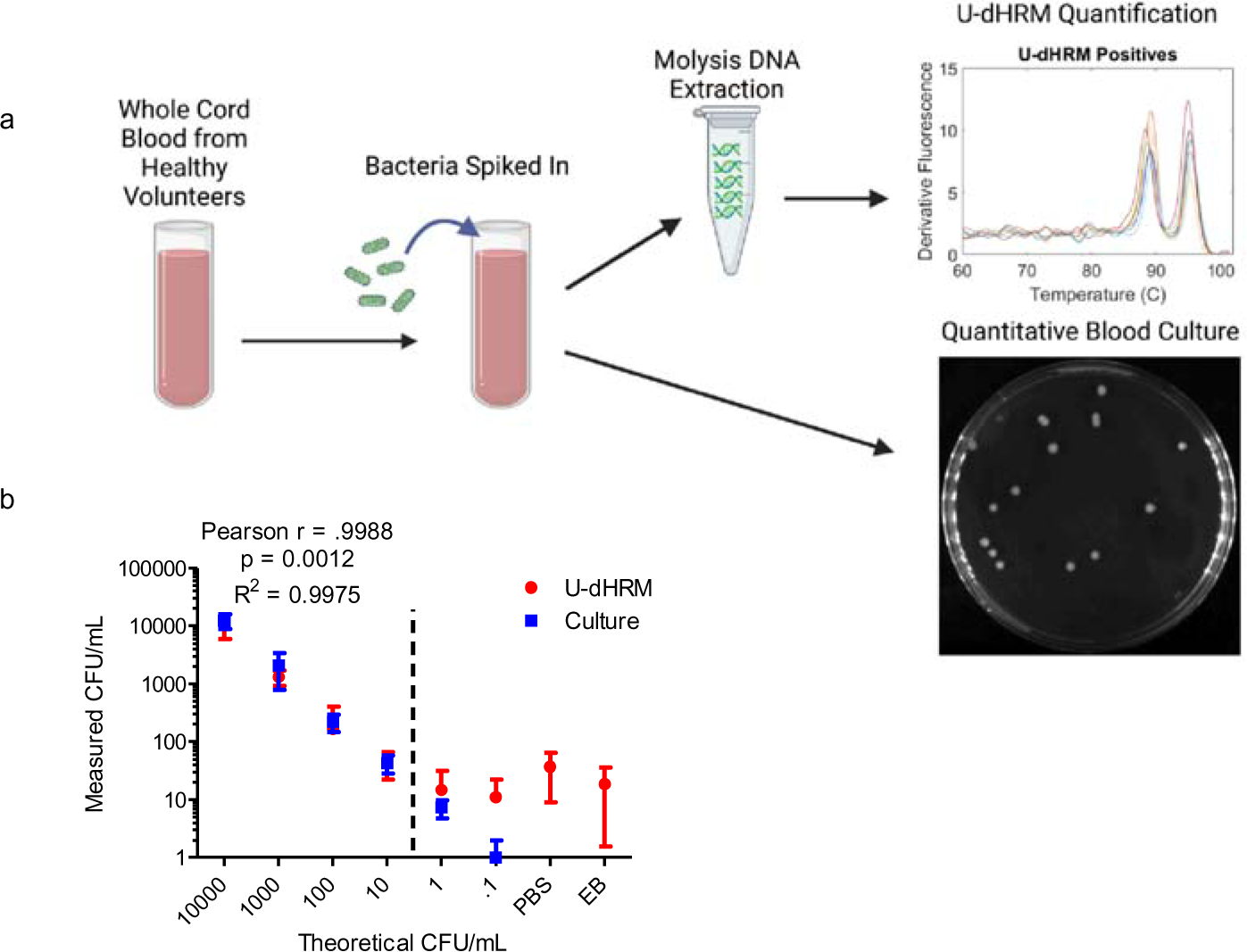
Analytical validation of U-dHRM on spiked whole blood samples. A) Schematic showing the procedure for analytical validation studies comparing U-dHRM to quantitative blood culture. B) Plot comparing quantification of bacteria by U-dHRM versus culture. A 6-fold dilution series of *E. coli* was prepared in PBS and spiked into whole blood. Controls included blank PBS spiked into blood and an extraction blank (EB, Molzyme SU buffer alone as input for Molysis processing). It is assumed that one 16S copy/melt curve will be detected per CFU.

### Pilot Clinical Testing

Next, we conducted U-dHRM analysis on 21 whole blood samples from patients ranging in age from infants to toddlers (Supplementary Table 3). These were remnants of complete blood count (CBCs) that were drawn from pediatric patients undergoing blood culture for suspicion of BSI. Only CBCs drawn at the same time from the same location as a blood culture draw were included (see Methods section). Of these 21 samples, 14 were matched to negative cultures and 7 were matched to positive cultures. Clinician adjudication of blood culture results was performed by a practicing physician in the Department of Pediatrics, Division of Infectious Diseases at Rady Children’s Hospital. The clinician reviewed the charts for each patient and each specific sample to determine if the blood culture results indicated a probable infection when compared to patient inflammatory markers, final diagnosis, additional diagnostic tests, success of prescribed treatment, and any other relevant clinical information. A summary of blood culture results, clinical diagnostic details for each sample, and clinician adjudicated diagnosis for blood culture positive samples is presented in Table 1.

**Table 1.** Comparison of clinical diagnostics and U-dHRM results.

| No. | Clinical TTR | Cultured Organism | ePlex BCID Result | Preliminary Diagnosis | Final Diagnosis | Clinician Adjudication | U-dHRM ID | U-dHRM Confidence Level |
| --- | --- | --- | --- | --- | --- | --- | --- | --- |
| 1 | 0d, 22hr, 44min | *S. epidermidis* (CoNS) | *S. epidermidis* (CoNS) | Acute UTI + Fever | *E. coli* UTI | Negative | CoNS | High |
| 2 | 2d, 12hr, 54min | *S. enterica* Typhi | *S. enterica* Typhi | Acute febrile illness in child | *S. enterica* Typhi BSI | Positive *S. enterica* | *S. enterica* | High |
| 3 | 3d, 0hrs, 45min | *S. enterica* Typhi | *S. enterica* Typhi | *Salmonella* BSI | *S. enterica* Typhi BSI | Positive *S. enterica* | *S. enterica*, *S. maltophilia* | High |
| 4 | 0d, 13hr, 24min | *S. maltophilia* | *S. maltophilia* | BSI | *S. maltophilia* CLABSI | Positive *S. maltophilia* | *S. maltophilia* | High |
| 5 | 1d, 12hr, 20min | *Salmonella* spp. | Negative | *Salmonella* BSI | *S. enterica* BSI | Positive *S. enterica* | *S. enterica*, *S. aureus* | High |
| 6 | 0d, 16hr, 59min | GPR | Bacteroides | Cellulitis | Poly-microbial SSTI | Negative | CoNS | High |
| 7 | 0d, 16hr, 36min | GNR | *Enterobacter non cloacae* | BSI | *Klebsiella* CLABSI | Positive *Klebsiella* | *K. pneumoniae* | Low |

\*CoNS = Coagulase Negative *Staphylococci*; BSI = Bloodstream Infection; UTI = Urinary Tract Infection; CLABSI = Central Line Associated BSI; SSTI = Skin and Soft Tissue Infection.

The machine learning algorithm, which was trained to classify database curves from spiked blood samples (Fig. 1), was used to automatically classify melt curves detected in the patient samples. Melt curves were determined to be either a high or low confidence match to the database organisms, or unmatched. Algorithms for high confidence, low confidence, and unmatched calling are detailed in the Methods and Supplemental Methods sections.

### Results for CBC Negative Samples

All of the 14 CBC samples matched to negative blood cultures were reported to be negative for any organism curves by the ML algorithm.

### Results for CBC Positive Samples

In positive CBC samples 1-5, ML curve classification identified high confidence matches to the same organisms identified by blood culture (Table 1). Representative melt curves detected by ML classification are shown in Fig. 3 A-I. In a few cases, additional organisms were detected by U-dHRM. In sample 3 where *S. enterica* was the primary pathogen detected by both BCID and U-dHRM (Fig. 3C), *S. maltophilia* was also detected at lower levels by U-dHRM (Fig. 3D). In sample 5, where *S. enterica* was detected by BCID and U-dHRM (Fig. 3F), *S. aureus* was also detected by U-dHRM (Fig. 3G).

**Figure 3.**
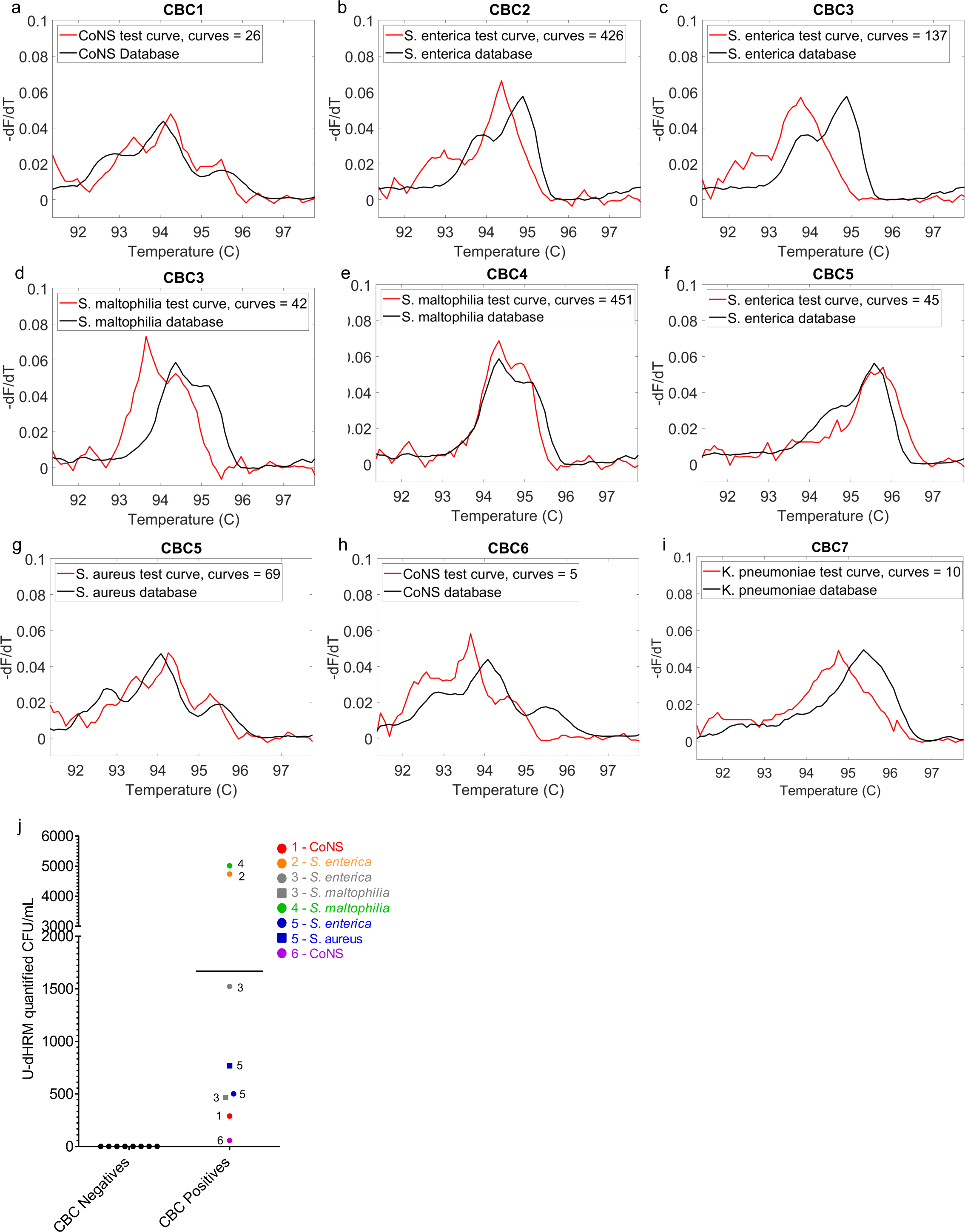
Pilot clinical testing on whole blood samples. A-I) Representative organism curves (red) that were matched with high confidence to database curves (black) by the machine learning algorithm in each CBC sample. No curves were detected in CBC negatives. The number of melt curves detected for each organism (cluster) is noted in the legends. J) Load of individual clinically relevant organisms identified and quantified by U-dHRM in CBC samples.

CBC samples 6 and 7 had discordant BCID and U-dHRM organism identifications. In sample 6, blood culture found gram positive rods (GPR) and post-culture ePlexID identified *Bacteroides*. Clinical adjudication determined this to be a false positive (Table 1). U-dHRM detected CoNS with high confidence (Fig. 3H).

In sample 7, blood culture found gram negative rods (GNR) and post-culture ePlexID identified *Enterobacter* non *cloacae*. However subsequent blood draws on this patient resulted in later detection of *Klebsiella* by BCID and clinician adjudication confirmed Klebsiella BSI (Table 1). U-dHRM did not identify high confidence matches in this sample, but did identify low confidence matches to *K. pneumoniae* (Fig. 3I). Low confidence matches for all samples are shown with their closest database matches in Supplementary Figures 8-11.

Fig. 3J shows the bacterial load quantified by U-dHRM in each CBC sample for high confidence matches. The time to result (TTR) for U-dHRM, 6hr, was substantially faster than BCID in all cases (Table 1). U-dHRM ranged from 7.5hr to 2d 18hr faster than BCID TTR for positives. Negatives are confirmed by BCID at 5d.

In summary, the overall concordance for the detection of bacteremia in whole blood samples was 21/21 (100%) from the perspective of blood culture as truth and 19/21 (90.5%) from the perspective of clinically adjudicated findings as truth. In the 5 samples determined to be true bacteremia by clinician adjudication, U-dHRM accurately identified the organism in 5/5, with 4 high confidence matches and 1 low confidence match.

## DISCUSSION

U-dHRM with ML achieved accurate organism identification in spiked whole blood samples and in clinical whole blood samples, suggesting that this approach can overcome a common pitfall of standard HRM where interference from biological substances in whole blood that co-extract with DNA cause large Tm shifts.^58^ U-dHRM overcomes this by generating thousands of example curves for each organism, which are identified independently of Tm by curve shape matching using ML. Interfering substances that carryover from blood can also impact the sensitivity and quantitative power of standard PCR-based assays by reducing amplification efficiency. U-dHRM achieved excellent quantitative agreement with QBC in the range of 10,000-10 CFU/mL in spiked blood by relying on endpoint detection, which overcomes efficiency bias and enables absolute quantification. Below 10 CFU/mL, U-dHRM was unable to accurately quantify because only ∼10% of each 1mL sample volume was tested. This limitation can be overcome by analyzing more of the extracted sample volume, concentrating the elute, running more chips per sample, increasing the number of digital partitions, or increasing the volume of digital partitions. Nonetheless, some differences between blood culture and molecular detection approaches may persist due to the presence of viable but non-culturable cells.^59^

In pilot clinical sample testing, U-dHRM demonstrated 14/14 (100%) agreement with culture negative samples. Given that universal bacterial primers form the basis of the assay, this result suggests that the testing process used was successful in limiting the detection of background and degraded DNA of no clinical significance. The sample preparation method used is specifically designed to bias detection towards DNA from intact/living bacteria cells by degrading cell-free DNA before microbe lysis. Also, the length of the V1-V9 amplicon, ∼1400 bp, biases towards non-degraded DNA.

U-dHRM also demonstrated 7/7 (100%) agreement with blood culture positivity, although two of these (samples 1 and 6) were determined by the adjudicating clinician to be false positive blood cultures. For sample 1, both BCID and U-dHRM detected CoNS. However, the final diagnosis was a urinary tract infection (UTI) caused by *E. coli*. The patient was not treated for an S. *epidermidis* BSI, but was treated for the UTI and improved, leading the adjudicating clinician to believe that the *S. epidermidis* was a contaminant. Since *S. epidermidis/*CoNS was detected by both methods, this was likely a true live cell contaminant. For sample 6, BCID detected *Bacteroides* while U-dHRM detected CoNS and two other curves that did not match any organisms in our database. The final diagnosis was polymicrobial skin and soft tissue infection (SSTI). In older children and adults, BCID of CoNS is usually considered to be a false positive caused by contamination, but in neonates it can indicate true BSI^60^. In the future, the absolute quantitative power of U-dHRM may be able to help to further refine these heuristics by defining clinically relevant load ranges that differentiate true bacteremia in need of treatment from common contamination levels for organisms that can be both pathogen and commensal.

The U-dHRM sample to answer time was 6hr for all samples, while BCID TTR ranged from ∼13.5hr to 3d for positive samples and up to 5d for negative samples. Interestingly, the positive samples with the longest TTR by blood culture (samples 2 and 3, TTR ∼2d and 3d respectively) were determined to have some of the highest bacterial loads by U-dHRM. Both U-dHRM and BCID identified *S. enterica*, which is known to grow slowly in blood culture because it is an intracellular pathogen that prefers to grow in macrophages.^61^ Blood culture and bone marrow culture are limited in their ability to recover this pathogen,^62^ even in patients harboring loads of 1.01*10^3^ to 4.35*10^4^ copies/mL.^63^ Clinical adjudication revealed that samples 2 and 3 originated from the same patient but were taken on subsequent days. U-dHRM quantified the first blood draw from this patient (sample 2) as having three times the concentration of *S. enterica* as the second draw on the following day (sample 3). However, U-dHRM also detected *S. maltophilia* in the second draw (sample 3) at a lower concentration than *S. enterica*, which may indicate a secondary infection in this patient since *S. maltophilia* is an opportunistic pathogen. The patient did not have antibiotic exposure at the time of either blood draw, so the decrease in *S. enterica* may have resulted from the patient naturally beginning to clear the infection. This highlights the possibility of using U-dHRM to track infections over time and in response to treatment.

*S. maltophilia* was also detected by U-dHRM and BCID in sample 4. *S. maltophilia* is an emerging pathogen of concern for nosocomial infections. It is being isolated more frequently and can be found ubiquitously in hospital environments.^64,65^ The time saved by U-dHRM detection of this organism (∼7hr) could have resulted in faster treatment with appropriate antimicrobial therapies, since this organism is inherently multidrug-resistant.^66^

In one case, sample 5, U-dHRM identified the causative pathogen while post-culture ePlex PCR failed. This sample took over a day to grow out in culture and be identified, meaning that U-dHRM would have provided a faster detection and identification time.

In sample 7, U-dHRM returned low confidence calls, with one of them partially matched to the causative pathogen, *Klebsiella.* Blood culture did not identify the causative pathogen in the matched sample. However, subsequent blood cultures did identify Klebsiella. Unfortunately, matched U-dHRM samples for these subsequent cultures were not available. Low confidence calls result from disagreement between the two ML classification methods used, which can arise due to the organism not being in the database or due to melt curve noise, which can prevent confident classification and identification. Expansion of the digital melt curve database with more organisms and more training curves for each organism is expected to improve curve calling confidence.

## CONCLUSIONS

Overall, the U-dHRM approach demonstrated 100% concordance with blood culture (positive/ negative) and 90.5% concordance with clinician adjudicated blood culture results (positive BSI/negative BSI). In all samples representing true bacteremia, U-dHRM correctly identified the causative pathogen, achieving 100% concordance with the genus and species identified by culture. U-dHRM has a much faster time to result than traditional blood culture methods, and automation of the U-dHRM process is expected to further reduce the sample-to-answer time. However, this pilot study of 21 patient samples needs to be further validated by testing a larger cohort across multiple medical centers. Additional organisms will also need to be added to the database to encompass all of the most clinically relevant organisms for sepsis. Further optimization of our ML algorithm and additional training data is expected to improve the ability of U-dHRM to call organisms with high confidence as well as to discern when samples contain organisms that are not yet in the database.

## ETHICS

The clinical study protocol was reviewed and approved by the Institutional Review Board (IRB) and Ethics Committee (No. 191392) of the University of California, San Diego (UCSD) and Rady Children’s Hospital (RCHSD). Consent was not required for the scavenging of remnant CBC samples.

## DATA AVAILABILITY

The data that support the findings of this study, in accordance with IRB designated restrictions, are available from the corresponding author upon request.

## ACKNOWLEDGEMENTS

This work was supported by the National Institute of Allergy and Infectious Diseases of the National Institutes of Health (award number R01AI134982), a Burroughs Wellcome Fund Career Award at the Scientific Interface (award number 1012027 to S.I.F.), and UCSD CTRI, FISP, and AIM pilot grants. We acknowledge and thank MelioLabs for the use of their MeltRead Platform and for their assistance in data analysis with their ML pipeline.

S.I.F. and A.A. designed the study. A.A. conducted experiments and analyzed data with assistance from T.G. N.R. provided clinical adjudication and general guidance in interpreting clinical results. S.M.L., K.M., and E.S. provided cord blood for analytical validation and database development and additional clinician guidance. A.K., A.S, and M.S. developed the algorithm used for machine learning and used it to perform quantification and classification, and assisted in interpretation of the machine learning results. D.P., P.K., K.L, and M.C. provided clinical isolates and performed blood culture testing on samples acquired from the blood bank. M.V., S.L, M.C, and Y.T. collected clinical samples and provided the associated clinical data from Rady Children’s Hospital.

## DISCLAIMER

The views expressed in this article are those of the author(s) and do not necessarily reflect the official policy or position of the Department of the Navy, Department of Defense, or the US Government.

