## Supplementary Materials for "Universal digital high resolution melt analysis for the diagnosis of bacteremia"

**Supplementary Figures**

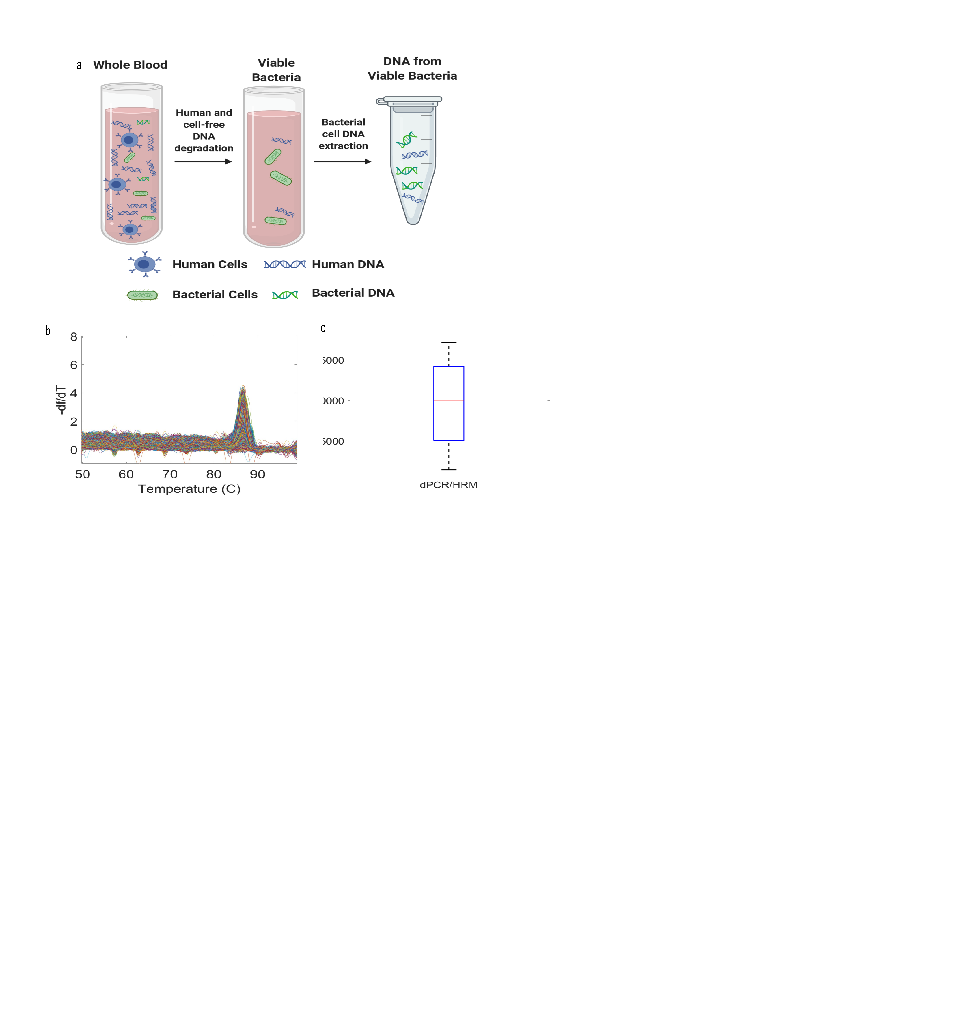

**Supplementary Figure 1. Quantification of human DNA concentration after Molysis processing of whole blood.** A) Schematic of the Molysis host DNA depletion and DNA extraction method. B) dHRM curves for β-actin gene amplification after Molysis processing. C) β-actin concentration remaining after Molysis processing as measured by dHRM across four replicates in two extractions.

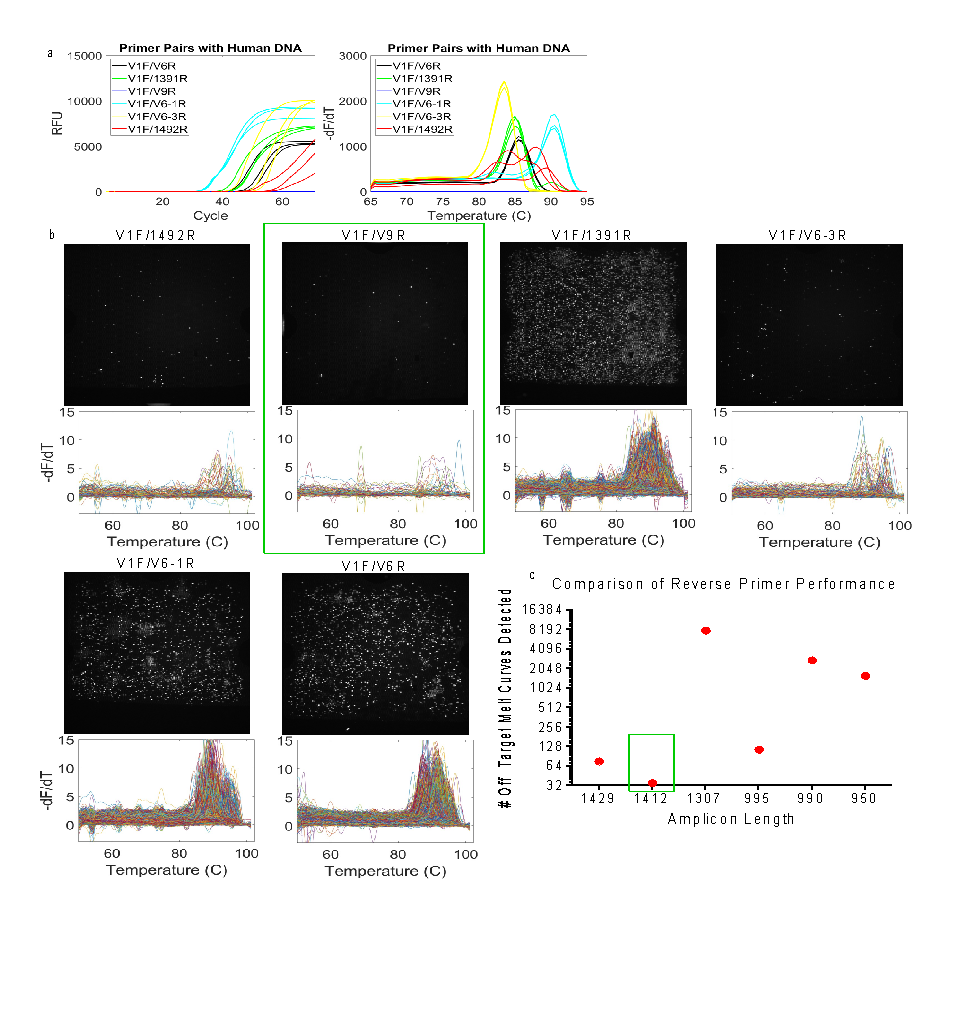

**Supplementary Figure 2. Universal bacterial primer screening for off-target amplification.** Six previously reported primer pairs were tested for off-target amplification of human DNA at the concentration of human DNA remaining after Molysis processing, which was determined in Fig. 1. A) qPCR amplification and HRM plots for all six primer pairs. B) Representative fluorescence images of dPCR chips (white indicates amplification positive partitions) and dHRM curves for each primer pair. C) Plot of number of melt curves detected for each primer pair versus the amplicon length. Green box denotes the best performing primer pair with the least amount of off-target curves, V1F/V9R.

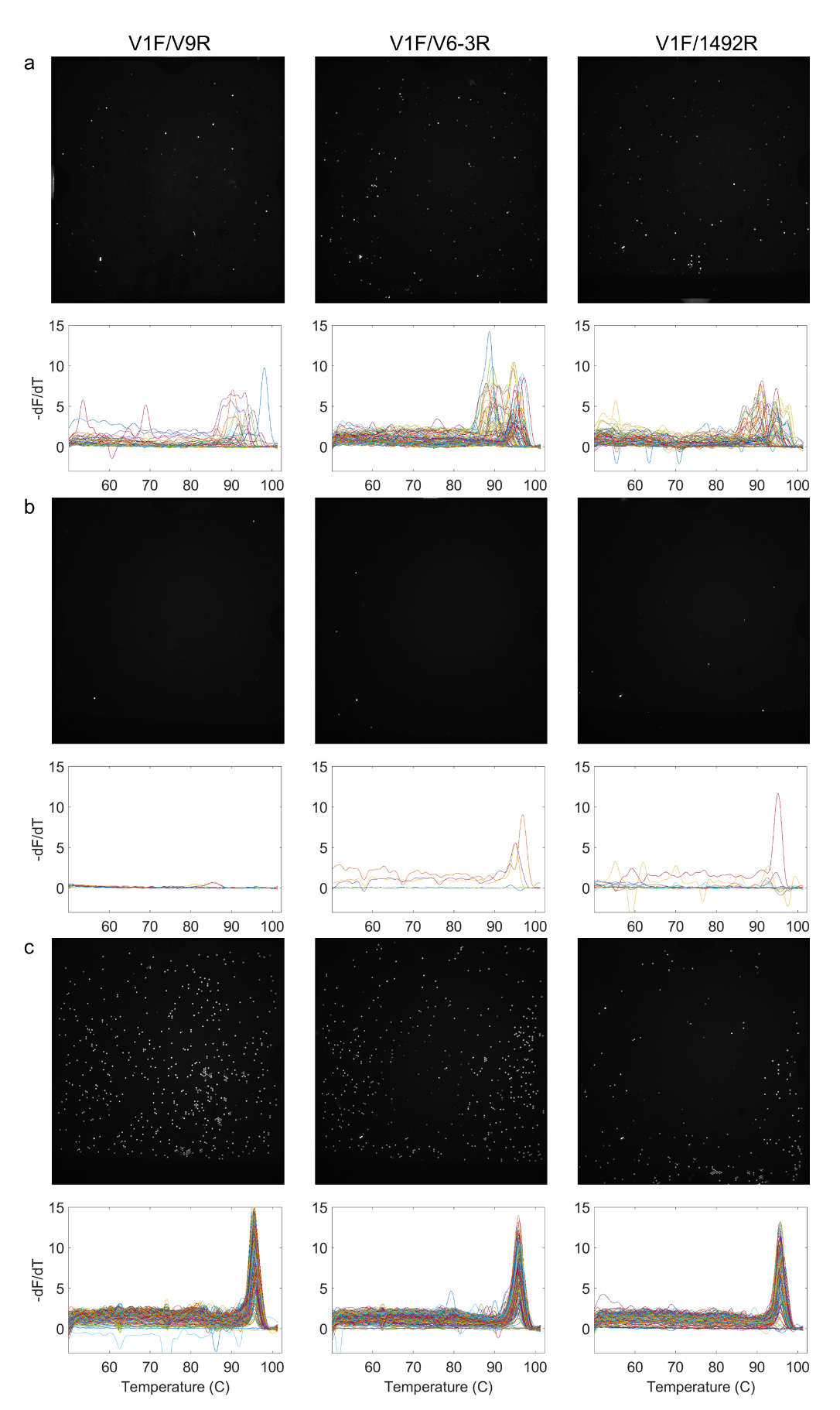

**Supplementary Figure 3. The three best performing primers from the first screening pass.** The following targets were amplified: A) human DNA, B) no DNA, C) bacterial DNA.

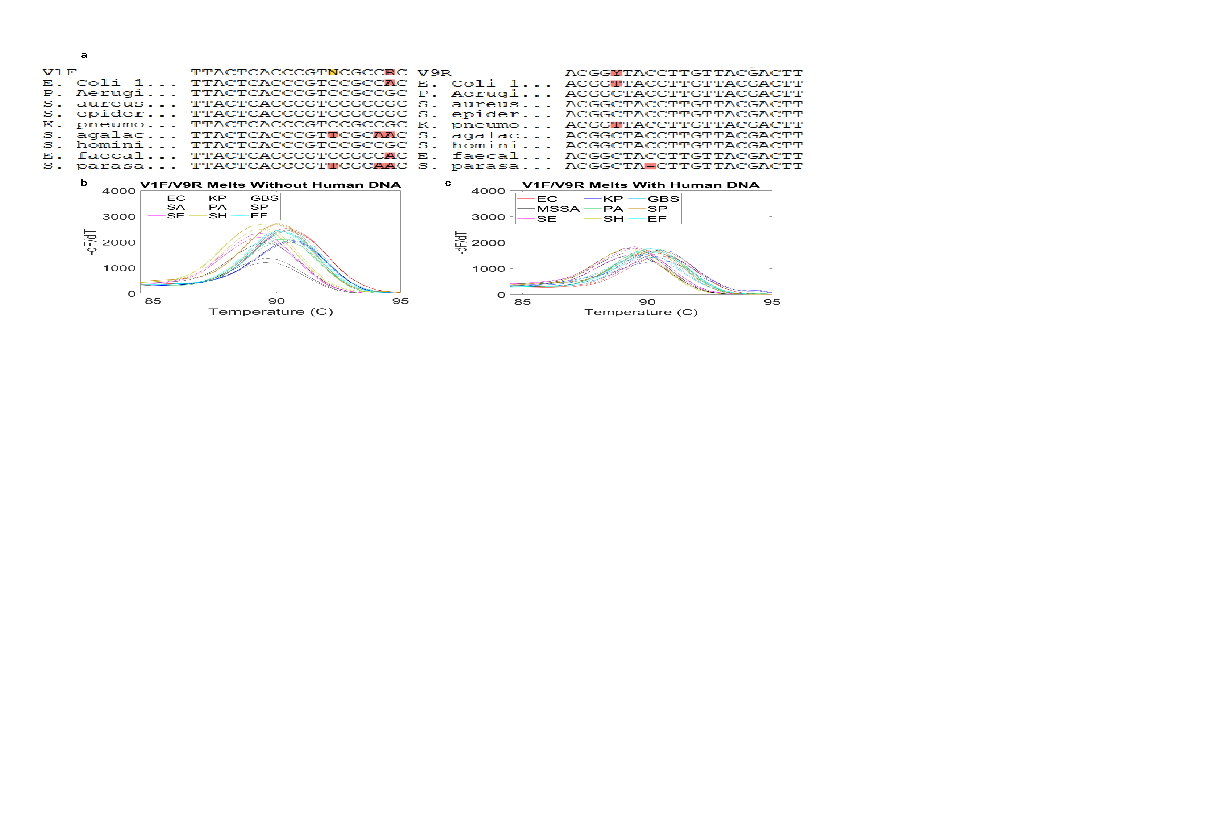

**Supplementary Figure 4. V1F/V9R universal bacterial primer screening for on-target amplification.** A) Alignment of best performing primer pair, V1F/V9R, and 16S rRNA gene sequences for nine bacterial pathogens known to be prevalent in San Diego ICUs. B) HRM curves for each pathogen target after qPCR (n=3) using V1F/V9R primers in the absence of human DNA and C) in the presence of human DNA at the post-Molysis concentrations determined in Supplementary Fig. 1.

   
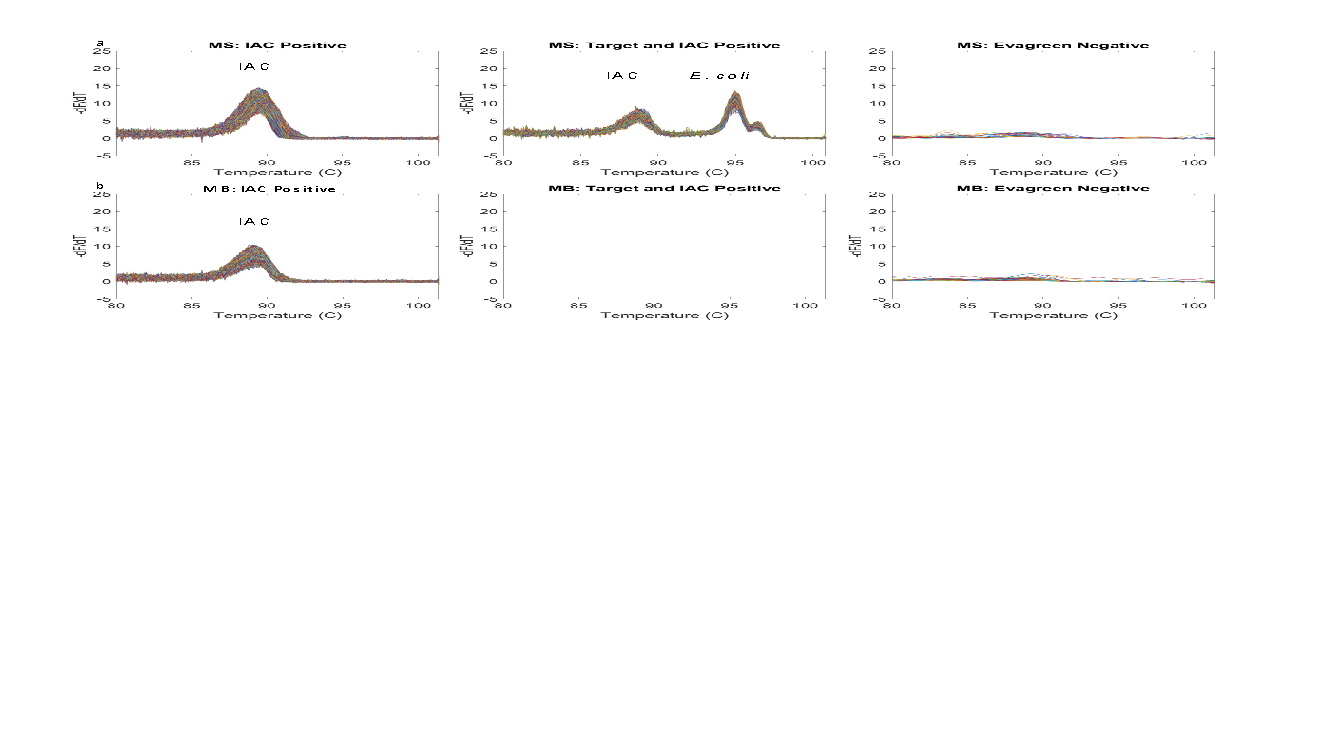

**Supplementary Figure 5. Validation of internal amplification control in matrix blank and *E. coli* matrix spike samples.** A) Representative dHRM curves in a matrix spike sample where partitions contain either IAC curves only (left), IAC and target curves (middle), or no curves (right, not capable of amplifying). B) Representative dHRM curves in a matrix blank sample where partitions with only IAC curves (left) or no curves (middle and right, not capable of amplifying) are expected.

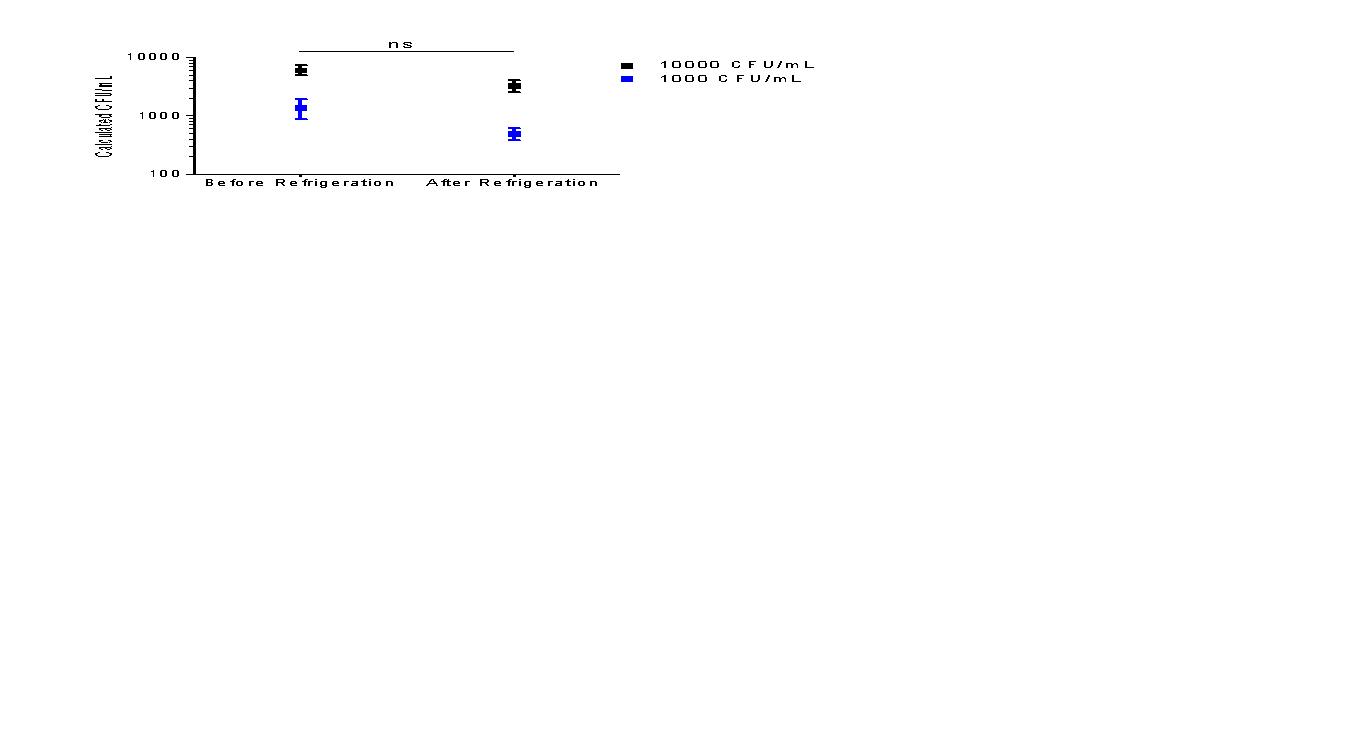

**Supplementary Figure 6. Mock Aging.** Mock aging studies were conducted to determine the change in quantification stemming from storage at 4°C on blood samples. Contrived infectious blood samples were created by spiking in two known concentrations of bacteria into healthy human cord blood and splitting the sample for immediate processing or for storage at 4°C for 24 hours before being processed by the Molysis DNA extraction method and subsequent U-dHRM analysis. The calculated concentration of bacteria in CFU/mL were compared before and after refrigeration.

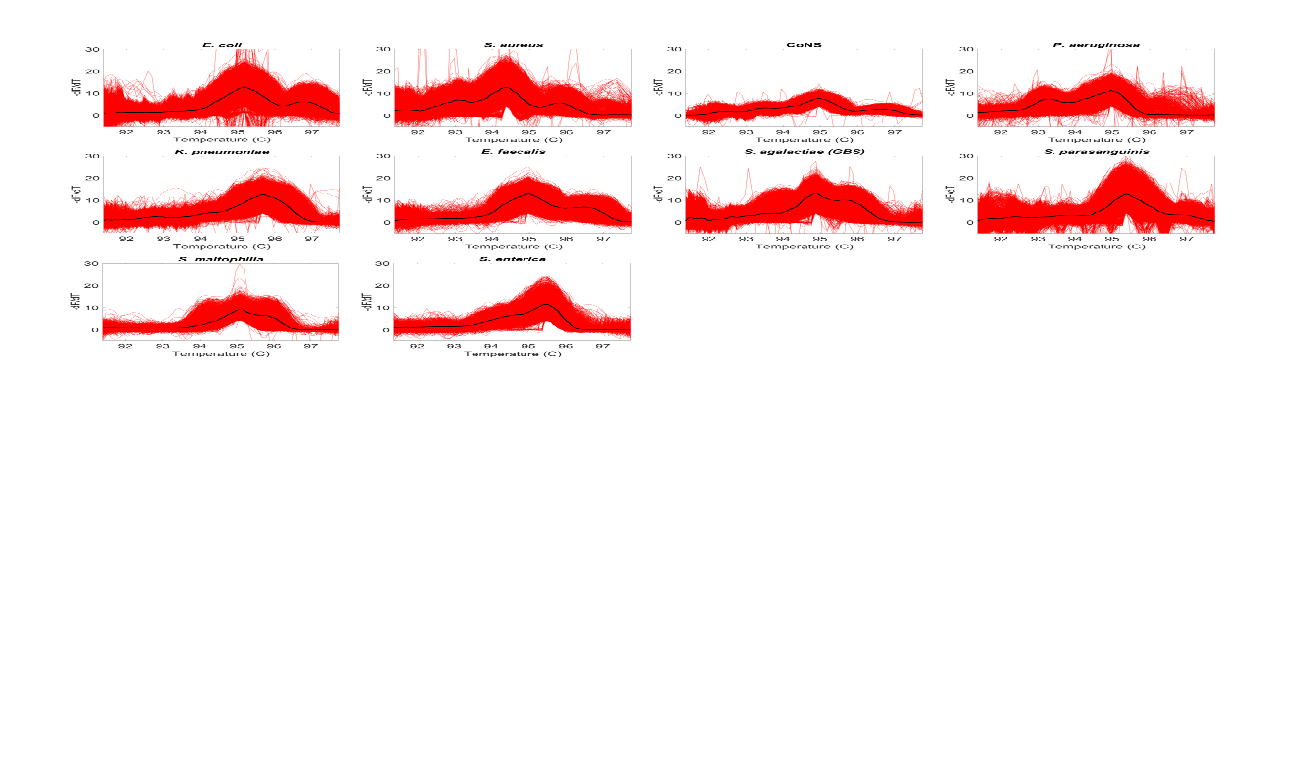

**Supplementary Figure 7. Raw database curves for 11 organisms.** All organisms are spiked into healthy human cord blood and pathogen DNA is acquired using the Molysis DNA extraction method. Following Molysis extraction, individual organisms were analyzed using U-dHRM. Raw dHRM curves are shown in red. The representative curve shapes identified by the ML algorithm for classification is shown in black. Since the ML algorithm was shown to have improved classification accuracy when CoNS is considered as a combined group, S. epidermidis and S. hominis are shown here as a combined CoNS group.

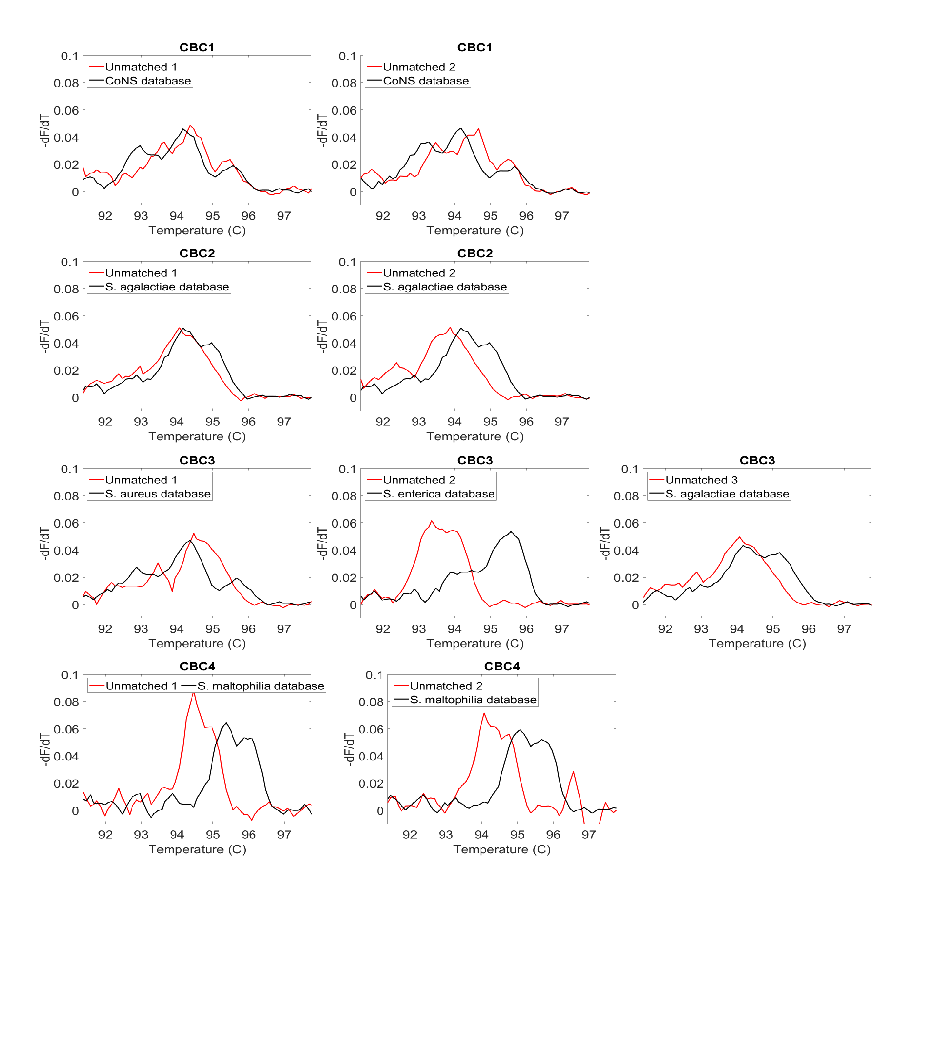

**Supplementary Figure 8. Novel and unmatched melt curves and their closest matching organism by DTW distance for samples 1-4.** Positively identified melt curves from each patient (red) overlaid with the matching database melt curve (black).

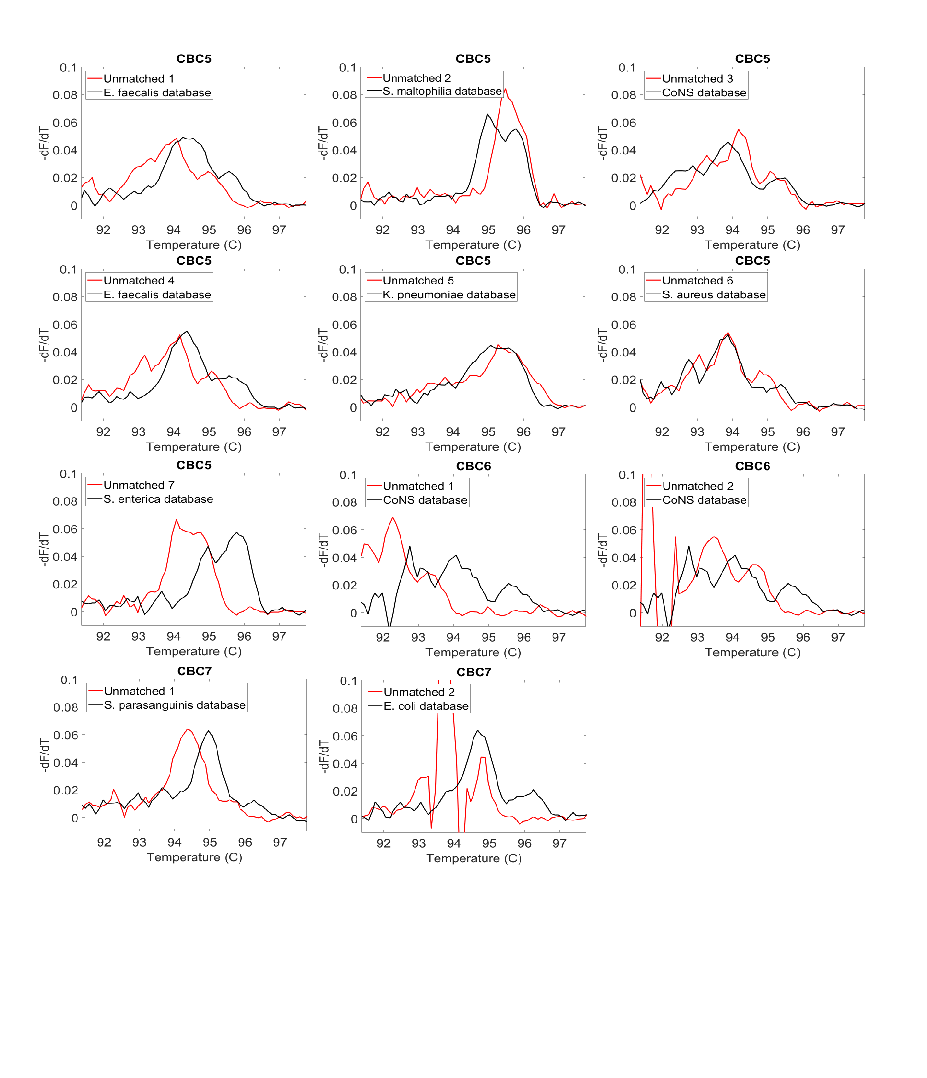

**Supplementary Figure 9. Novel and unmatched melt curves and their closest matching organism by DTW distance for samples 5-7.** Positively identified melt curves from each patient (red) overlaid with the matching database melt curve (black).

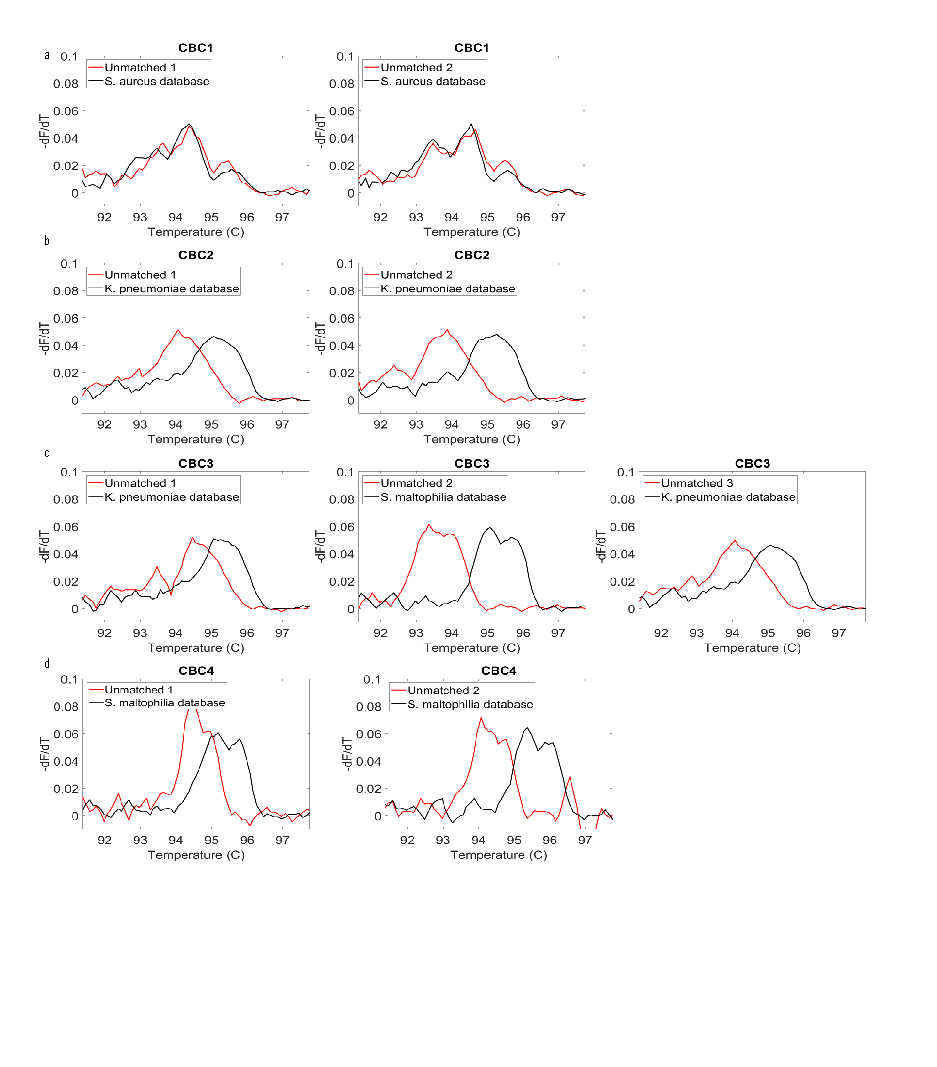

**Supplementary Figure 10. Novel and unmatched melt curves and their closest matching organism by Euclidean distance for samples 1-4.** Positively identified melt curves from each patient (red) overlaid with the matching database melt curve (black).

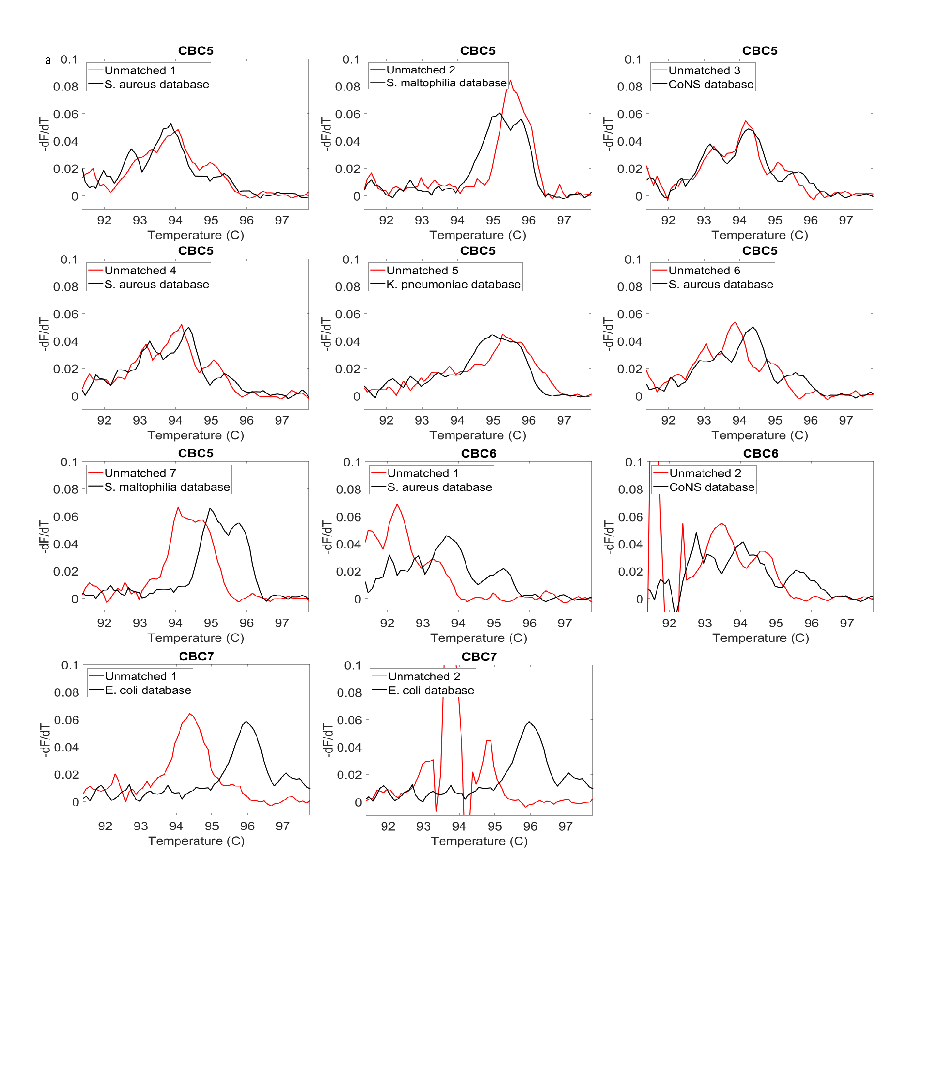

**Supplementary Figure 11. Novel and unmatched melt curves and their closest matching organism by Euclidean distance for samples 5-7.** Positively identified melt curves from each patient (red) overlaid with the matching database melt curve (black).

**Supplementary Tables**

**Supplementary Table 1. Primers screened for universal 16S amplification and off-target interactions with human DNA.**

| **Primer Name** | **Sequence (5’-3’)** | **16S Amplicon Length** | **Human Query Coverage** | **Human % Identity** | **Reference** |
| --- | --- | --- | --- | --- | --- |
| V1F | GYGGCGNACGGGTGAGTAA | - | 63% | 100% | ^1^ |
| V6R | AGCTGACGACANCCATGCA | 950 | 82% | 100% | ^1^ |
| V6_1R | ACGAGCTGACGACARCCATG | 990 | 60% | 100% | ^2^ |
| V6_3R | ACAACACGAGCTGACGAC | 995 | 100% | 89.47% | ^3^ |
| 1391R | GACGGGCGGTGTGTNCA | 1307 | 77% | 100% | ^4^ |
| V9R | ACGGYTACCTTGTTACGACTT | 1412 | 70% | 100% | ^5^ |
| 1492R | CGGTTACCTTGTTACGACTT | 1429 | 61% | 100% | ^6^ |

**Supplementary Table 2. Eleven bacterial species selected for screening.**

| **Organism** | **Culture Media** | **Strain ID** |
| --- | --- | --- |
| *E. coli* | Luria Bertani | NBRC 102203 |
| *S. aureus* | Tryptic Soy | ATCC BAA-1721^6^ |
| *S. epidermidis* | Tryptic Soy | ATCC 700576 |
| *P. aeruginosa* | Nutrient Broth | Clinical Isolate |
| *K. pneumoniae* | Tryptic Soy | ATCC BAA-1705 |
| *S. hominis* | Nutrient Broth | ATCC 27845 |
| Group B Streptococcus *(S. agalactiae)* | Todd Hewitt | Clinical Isolate |
| *S. parasanguinis* | Brain Heart Infusion | Clinical Isolate |
| *E. faecalis* | Brain Heart Infusion | Clinical Isolate |
| *S. enterica* | Tryptic Soy | Clinical Isolate |
| *S. maltophilia* | Tryptic Soy | Clinical Isolate |

**Supplementary Table 3. Age range definitions for patient samples.**

| Age Range Definitions | | |
| --- | --- | --- |
| Infant | CBC 1 | 29 days - 1 year |
| Toddler | CBC 5, 6, 7 | >1, <5 years |
| Child | CBC 2, 3, 4 | >5, <13 years |
| Teenager |  | 13-18 years |

**Supplementary Table 4. Cross validation results for database curves**

| **Organism** | **Precision** | **Recall** | **F1-Score** | **Support** |
| --- | --- | --- | --- | --- |
| CoNS  (*S. epidermidis, S. hominis*) | 0.86 | 0.91 | 0.88 | 10870 |
| *E. coli* | 0.97 | 0.96 | 0.97 | 18845 |
| *E. faecalis* | 0.94 | 0.95 | 0.94 | 10185 |
| *K. pneumoniae* | 0.97 | 0.97 | 0.97 | 10987 |
| *S. aureus* | 0.95 | 0.93 | 0.94 | 20450 |
| *P. aeruginosa* | 1.00 | 0.99 | 0.99 | 11482 |
| Group B Strep (*S. agalactiae*) | 0.99 | 0.98 | 0.99 | 26422 |
| *S. enterica* | 0.98 | 0.99 | 0.98 | 12119 |
| *S. maltophilia* | 0.99 | 0.98 | 0.98 | 3394 |
| *S. parasanguinis* | 0.98 | 0.99 | 0.99 | 21550 |
| accuracy |  |  | 0.97 | 146304 |
| macro avg | 0.96 | 0.96 | 0.96 | 146304 |
| weighted avg | 0.97 | 0.97 | 0.97 | 146304 |

**Supplementary Methods**

***Data Curation & Preprocessing:***

To generate the melt curves, we followed a similar protocol to the one previously published for fluorescence data extraction ^7^. Each melt curve is a sequentially ordered dataset measured at regular time intervals and can be treated as a time series ^8^. As detailed in our previous publication, fluorescence values are recorded at the rate of two fluorescence images per second (2 Hz) during post-amplification chip-heating at 0.2 °C / sec ramp rate from 50 °C to 103 °C ^7^. The melt curve is a time series with fluorescence values recorded every 0.5 sec time interval, equivalent to 0.1 °C interval on the temperature axis.

We take the derivative of the raw fluorescent melt curves for further analysis. We utilize the *gradient()* function of the *numpy* python package to obtain the derivatives from the melt curves ^9^.Melt curve derivatives with internal control and bacterial peaks were labeled positive. To identify the peaks, we utilized the *find_peak()* function of the *scipy* python package ^10^. Peak detection was performed above and below a temperature of 96°C, for the bacterial and internal control peaks respectively. When both peaks were detected, we considered it a positive melt curve. We utilized these positive melt curves to train the classifier. In contrast, the negatives, which correspond to melt curves having only one peak or no peaks at all, were excluded from consideration. Following preprocessing, the following steps were applied to each positive melt curve. First, the Savitzky-Golay (SG) filter with window length nine (equivalent to 4.5 sec time interval or 0.9 °C temperature change) and polynomial order three was applied ^11^. These parameters of the SG filter were chosen based on visually inspecting the fit quality. For example, using a lower polynomial order or a wider window length can result in excessive smoothing, leading to the loss of the original shape. After that, each melt curve was further sliced to retain only the bacterial region. Lastly, the Area Under the Curve (AUC) normalization was applied to them.

***Melt Curve Classification:***

Distortion (or shift) along the temperature (or time) axis causing well-to-well as well as chip-to-chip variations in melt curves is something inherent in HRM ^12–14^. From a classification perspective, this means that one needs a classifier that classifies two melt curves into the same class when they are similar but slightly shifted along the time (or temperature) axis. Temporal distortions of such kind can be dealt with by using the various elastic distance measures for time series such as Dynamic Time Warping (DTW) and its variations ^15–17^. Intuitively, DTW is a non-linear alignment technique allowing one-to-many alignment of points between pairs of time series. A distance matrix is constructed between two time series T and T’ such that each element (i, j) in this matrix is the Euclidean distance between the i^th^ point of T and the j^th^ point of T’. Dynamic programming based methodology is then employed on this matrix to find a series of adjacent cells that begins and ends at diagonally opposite corners (warping path) and is minimal – called the minimal warping path. The length of this minimal warping path is the DTW distance between the two time series. For more information on DTW and its variants, we refer the readers to the extensive survey by Bagnall et. al. ^18^. DTW has also been used for both classification and noise modeling of HRM curves ^14,19^.

We leverage the elasticity of DTW by using it in a two-step classification process. In the first step, the melt curves for each organism are clustered, and in the second step a distance-based k-Nearest Neighbour (kNN) classifier is used to classify a test curve by comparing it with the cluster centers (obtained from the first step). As the melt curves have significant variations and noise, the clustering step helps in extracting the key clusters of variations. The cluster centers are effectively compressed signals that are more robust than the actual noisy melt curves and summarize the key patterns available in the database for a particular organism. Further, in the second step, we have to compare the test curve to only the cluster centers rather than all the database curves, thus providing efficiency benefits.

Specifically, 10% of the curves were randomly selected (*training data*) for each organism and then clustered using the well-known K-mean clustering technique^20^. In K-means, cluster assignment is first performed for each data point (based on their distance from the cluster center), and then averages of each cluster’s points are assigned as the new cluster center. Starting with random cluster centers, these two steps are repeated until no further changes in cluster membership. However, instead of using the usual Euclidean distance-based K-means, we use DTW for both the cluster assignment as well as the averaging step of K-means. More specifically, when DTW distance is used, a more suitable DTW-based Barycenter Averaging (DBA) technique proposed by François et. al for the K-means averaging step is utilized ^21^. The average sequence (cluster center) returned by DBA-based K-means for each cluster is called the *barycenter*. A set of barycenters is created for each organism from the clustering step. More specifically, we have *p* species and *k_i_* number of clusters for species *i* ∈ {*1, 2,* … , *p*} and the set of *k_i_* barycenters *B_i_* = {*b_1_, b_2_, …, b_k_i_*} obtained after clustering training data for species *i.* We then have the set of all the barycenters across the species as *B* = {*B_1_* ∪ *B_2_* ∪ …. ∪ *B_p_*}.

For the second step, we use Dynamic Time Warping (DTW) based kNN to classify a test curve from *test data* (the remaining 90% of curves in the database). More specifically, each curve *c* from the test data was compared against each barycenter *b* ∈ *B*, and the DTW distance, *dtw(c, b)*, for the same, was recorded. For example, we consider *B_topk_* to be the subset of *B (B_topk_* ⊆ *B)* containing the *k* nearest barycenters to test curve *c*. A test curve *c* is then classified as the organism whose barycenters are the majority of the set *B_topk_*. If there is no clear majority, we simply keep the test curve *c* as inconclusive. We refer to the selected species as *i_dtw_**. We also observed that DTW in some scenarios matched significantly distorted curves. To control these cases, we also perform a final check as follows. We find the barycenter closest to the test curve *c* in terms of Euclidean distance and check if the species identified by this barycenter agrees with the *i_dtw_** (the species identified using DTW-based kNN previously). The Euclidean distance is less elastic, and serves as a control to limit classification inaccuracies stemming from the elasticity of DTW. The combination of both types of distance measurements provides confident classification of each test curve. If there is a disagreement between the species (classes) identified by the DTW and Euclidean, we again tag the test curve *c* as inconclusive.

Here we describe the parameter selection and implementation details. K-means clustering requires the number of clusters (*k_i_* for each species *i*) as its input, and we used the well-known *elbow method* to find the number of clusters^22^. For kNN, the value for the number of nearest neighbors (or barycenters) was chosen as *k=3* after 5-fold cross-validation from the set *k* ∈ {*1, 3,* *5, 7*, *9*}. For both DTW calculations for kNN and K-means clustering using DTW, we used the *tslearn* python package. Specifically, we used the *dtw()* function (in the *metrics* subpackage of *tslearn*) for calculating DTW distances. For clustering, we used *TimeSeriesKMeans()* function (inside the *clustering* subpackage), which has the option of choosing DTW as distance metric for clustering.

**Clinical Validation:**

Similar preprocessing was done on the patient/clinical data to extract the positive melt curves and slice out the region of interest. We also excluded melt curves from wells that were spatially clustered. For the clinical melt curves obtained from a chip, we identified one or more curve subgroups of a significant size which could be classified into distinct bacterial species. The clinical curves are also prone to significant variations and noise, so we first performed clustering using DBA-based K-means. The barycenters thus obtained from the clinical data are then classified using the DTW-based kNN classifier (along with the Euclidean distance filter) built using the database melt curves (described in the previous subsection). Additionally, for a given clinical barycenter, *b^c^*, we consider only that database barycenter, *b_i_ ∈ B*, as a neighbor (for kNN classification) for which the DTW distance, *dtw(b^c^, b_i_)* is less than the maximum DTW distance of database barycenter, *b_i_* , to any other curve in its own cluster (i.e. database cluster corresponding to *b_i_*). If there is no such *b_i_* , then *b^c^* is tagged as *unmatched*. These *unmatched* curves can be potentially *novel* curves that are not yet present in our database.
